# Recent COVID-19 vaccination is associated with modest increases in the physiological demands to graded exercise

**DOI:** 10.1101/2021.08.24.21262239

**Authors:** Helena Batatinha, Forrest L. Baker, Kyle A. Smith, Tiffany M. Zúñiga, Charles R. Pedlar, Shane C. Burgess, Emmanuel Katsanis, Richard J Simpson

## Abstract

Athletes are advised to receive the COVID-19 vaccination to protect them from SARS CoV-2 infection during major competitions. Despite this, many athletes are reluctant to get the COVID-19 vaccine due to concerns that symptoms of vaccinosis may impair athletic performance.

**OBJECTIVE:** To determine the effects of COVID-19 vaccination on the physiological responses to graded exercise.

**METHODS:** Healthy participants completed a 20-minute bout of graded cycling exercise before and ~21 days after COVID-19 vaccination (2 dose Pfizer mRNA or 1 dose Johnson & Johnson).

**RESULTS:** Oxygen uptake, CO_2_ production, respiratory exchange ratio, ventilation, heart rate, serum noradrenaline, and rating of perceived exertion were significantly elevated in the post vaccine trial. However, vaccination did not affect serum lactate, adrenaline, cortisol, predicted 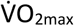, and ventilatory threshold. Post-vaccine effects on heart rate and noradrenaline remained significant in non-infected participants that received the Pfizer vaccine. No significant effects in respiratory gas exchange parameters were found after vaccination in those previously exposed to SARS-CoV-2, but exercise adrenaline levels were significantly lower and serum lactate levels trending (p= 0.10) lower after vaccination. No changes in any physiological responses to exercise were found in control participants who completed two bouts of exercise separated by ~5 weeks without vaccination.

**CONCLUSION:** Recent COVID-19 vaccination is associated with modest increases in the physiological demands to graded exercise in non-infected healthy people but may actually improve metabolic responses to exercise in those previously infected with SARS-CoV-2. Whether or not these small effects could impact athletic performance at the elite level warrants investigation.

## Introduction

The severe acute respiratory syndrome coronavirus 2 (SARS–CoV-2) - etiological agent of coronavirus disease 2019 (COVID-19)- was first identified in December 2019 in Wuhan, China, before being declared a global pandemic in March 2020 (1). As of August 2021, more than 200 million people worldwide have been infected with ~4.3 million deaths. The rapid production and distribution of mRNA (e.g. Pfizer and Moderna) and viral vector-based vaccines (e.g. Johnson&Johnson and AstraZeneca) that started in November 2020 has greatly limited the spread of COVID-19 (2), with 15% of the world’s population now fully vaccinated (3). Several clinical trials have demonstrated the safety and efficacy of the current COVID-19 vaccines (4), with reported side-effects such as body aches, fever, arm soreness, malaise and flu-like symptoms usually mild and resolving within 48h (5). However, reports are emerging that COVID-19 vaccination in a minority of patients has been associated with more severe and longer lasting symptoms including myocarditis fatigue, shortness of breath, cough, joint and chest pain (6).

It is generally recommended that elite athletes receive all necessary vaccines prior to competition due to increased risks of viral exposure (7). A recent study in elite German athletes of 10 different types of sport found that the quadrivalent inactivated influenza vaccine (a synthetic vaccine consisting of four inactivated influenza viruses) elicited a strong immune response one week after vaccination with no reported side-effects or loss of training (8). However, due to increasing reports (albeit mostly anecdotal) of adverse symptoms associated with COVID-19 vaccines, there is a growing concern among the athletic community that vaccination might hinder athletic performance. Such concerns could result in many athletes refusing to get vaccinated prior to competition and, consequently, contracting SARS-CoV-2 during upcoming major sporting events. Indeed, during current/recent sporting events such as the 2021 European Championship and Copa America international soccer tournaments, as well as the Tokyo Olympic Games, there have been several reports of players/athletes having to miss games/competition due to contracting SARS-CoV-2 or having been in contact with infected individuals.

To this end, there is a critical need to determine if COVID-19 vaccination impacts physiological responses to exercise to either alleviate or confirm concerns that athletes may have regarding potential negative effects of recent COVID-19 vaccination on athletic performance. Moreover, whether exercise responses are affected differently between mRNA and viral-vector based vaccines, as well as in those previously infected with SARS-Co-V2, needs to be determined. Here we investigated the effects of recent (within 2-3 weeks) COVID-19 vaccination on metabolic and physiological responses to graded cycling exercise in healthy individuals. We found modest increases in heart rate (HR), ventilation and serum noradrenaline levels during vigorous intensity exercise after vaccination. While these small physiological changes are unlikely to negatively affect exercise performance in the vast majority of healthy people, whether or not they could impact athletic performance at the elite level warrants investigation.

## Methods

### Participants

A total of eighteen (9 females, 9 males) healthy individuals between the ages of 24-43 years participated in this study. Baseline anthropometric and cardiovascular characteristics are shown in Table 1. Twelve participants received a COVID-19 vaccine during the study period (Pfizer mRNA vaccine (n=9), Johnson&Johnson viral vector-based vaccine (n=3)) while six participants, who were involved in a parallel non-vaccine related research study in our laboratory, served as controls. Prior to their enrollment, each subject completed an AHA-ACSM preparticipation screening questionnaire and medical history survey (9) to verify that they had not been previously diagnosed with any cardiovascular, metabolic, renal, liver, pulmonary, asthmatic, rheumatic, or other inflammatory disease/condition and were not currently under the administration of medication known to alter their inflammatory or metabolic profiles. All participants were additionally screened for physical activity participation to ensure the enrollment of non-sedentary individuals – physical activity rating, minimal score = 4 (10). Moreover, research participants were non-users of tobacco products and consumed ten or less standard alcoholic beverages per week on average. Participants were asked to abstain from alcohol, caffeine, and physical activity 24h prior to exercise trials and complete an overnight (minimum 8h and maximum 12h) fast prior to each laboratory visit. Adherence to these pre-testing procedures were confirmed verbally with the participants upon their arrival to the laboratory. All participants provided written informed consent prior to participating in the study and the Institutional Review Board (IRB) of the University of Arizona granted ethical approval.

**Table 1.** Participant demographic data presented as median ± SD

|  | **All (n=18)** | **Non-Infected (n=8)** | **Infected (n=4)** | **Controls (n=6)** |
| --- | --- | --- | --- | --- |
| **Female** | 9/18 | 5/8 | 1/4 | 3/6 |
| **Age (yrs)** | 29.5 ± 5.4 | 29 ± 3.9 | 35 ± 4.3 | 28 ± 8.4 |
| **Height (cm)** | 171.6 ± 11 | 170.5 ± 11 | 175.3 ± 10.4 | 177.5 ± 11.7 |
| **Weight (kg)** | 66.3 ± 13.6 | 65.1 ± 10.9 | 67.9 ± 19.3 | 70.2 ± 11.5 |
| **Resting HR (bpm)** | 70 ± 5.7 | 70 ± 5.6 | 69 ± 5.6 | 70 ± 5.9 |
| **Resting Systolic Blood Pressure (mmHg)** | 116 ± 8.2 | 115 ± 6.4 | 120 ± 12.2 | 119 ± 7.9 |
| **Resting Diastolic Blood Pressure (mmHg)** | 77 ± 6.7 | 76 ± 5.5 | 78 ± 9.4 | 77 ± 4 |
| **Predicted VO<sub>2max</sub> (mL/kg/min)** | 40.8 ± 9.9 | 42.5 ± 7.2 | 36.3 ± 14.1 | 44.1 ± 8.1 |
| **Time between main exercise trials (days)** | 52.5 ± 21.6 | 49 ± 15.7 | 70 ± 32.2 | 26 ± 185.2 |
| **Time between final vaccine dose and last exercise trial (days)** | 14 ± 10.1 | 13 ± 6.5 | 26 ± 12 | N/A |

### Experimental Design

The study required participants to visit our laboratory on three separate occasions. Visit 1 involved a pre-screening procedure to verify that participants were eligible for the study and healthy enough to perform vigorous intensity exercise and to provide written consent (ACSM/AHA questionnaire). If eligible participants completed a submaximal graded exercise test on a cycling ergometer (Velotron, Quarq Technology, San Diego, CA) to determine predicted maximal oxygen consumption 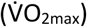. Blood samples were also collected during this visit to confirm serological status against SARS-CoV-2 using a commercially available ELISA kit (SARS-CoV-2 Spike S1 Human IgG; Biolegend, San Diego, USA). Visit 2 occurred 1-3 weeks after the first visit and required the participants to complete a continuous 20-minute graded cycling exercise with multiple blood collections from an intravenous catheter. Visit 3 required participants to perform the exact same trial that was performed during Visit 2 at 1-3 weeks after receiving the final COVID-19 vaccine dose via their own health care provider. This corresponded to an elapsed time of 5-7 weeks between Visit 2 and Visit 3. Participants arrived at our laboratory at the exact same time of day across all trials, which were performed between 06:00-09:00 local time.

### Submaximal Exercise Testing Procedure (Visit 1)

Upon arrival at the laboratory, participants were briefed regarding the nature of the testing protocol, and height, weight and resting blood pressure measurements were collected. Each participant was assessed for appropriate apparatus sizing (e.g., metabolic cart face mask) and cycling ergonomics (e.g., saddle height, handlebar reach, etc.) and these were recorded so they could be replicated during subsequent visits. Prior to initiating the test, all participants performed 3-5 minutes of seated rest on the cycling ergometer) for the collection of resting heart rate and respiratory gas exchange data. This was followed by a 5-minute warm-up period at 50 watts (W). Thereafter resistance was increased by 15 watts every minute and participants were asked to maintain a consistent cycling cadence throughout the entire exercise bout (≥60rpm). Exercise continued until the participant reached 85% of age-predicted maximum heart rate (220-age). Estimated 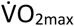 was determined using the built-in algorithm contained within the metabolic cart software (Quark CPET, COSMED, Pabona di Albona Laziale, Italy). Heart rate and rating of perceived exertion (RPE; Modified BORG 0-10 scale - (11)) were recorded during the final 15 seconds of every exercise stage. Individual linear regression equations were established for each participant and used to determine cycling power outputs corresponding to various percentages of the 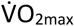 for the main exercise trials performed during Visit 2 and Visit 3.

### Main Exercise Trial (Visit 2 and 3)

During Visit 2 and Visit 3, participants’ weight was re-recorded, and they were fitted with an indwelling intravenous catheter (BD, Franklin Lakes, NJ, USA) inside an antecubital vein so that serial blood draws could be collected before, during and after exercise. After each blood draw, the catheter was flushed with isotonic saline and a 2mL was drawn from the catheter and discarded prior to collecting the blood sample used for analysis. Blood was collected into a 6mL vacuum tube containing a serum separator gel (BD Vacutainer® blood collection tubes). Participants were then asked to complete a 5-minute warm up at 50W before cycling continuously for an additional 20-minutes at graded intensities. The 20-minute trial consisted of four incremental 5-minute stages with power outputs corresponding to 50%, 60%, 70%, and 80% of the individual predicted 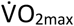. Participants again were asked to maintain a consistent cycling cadence throughout the entire exercise session (≥60rpm) and heart rate and respiratory gas exchange were measured throughout with RPE being recorded during the final 15 seconds of each exercise stage. Blood samples were collected at 4 separate time points during these visits: (1) at rest; (2) during the 60% 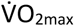 stage; (3) during the 80% 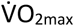 stage; and (4) at 1h after exercise cessation. An exception to this was the control participants who performed identical exercise protocols as part of a parallel but separate research study in our laboratory but had blood collected at rest and during the 80% 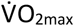 stage only. To maintain consistency, the absolute cycling power outputs for each individual were identical during Visit 2 and Visit 3. During Visit 3, the resting serum sample was also used to confirm that all vaccinated individuals had seroconverted and presented with a positive SARS-CoV-2 IgG titer.

### Assessment of Serum Biomarkers

Blood collected into vacutainers containing a serum gel separator were allowed to rest for 30 minutes and subsequently centrifuged at 1500 RCF for 10 minutes. Serum was then collected and stored at −80°C until future analysis of cortisol (EIAHCOR, INVITROGEN^®^, Frederick, MD, USA), lactate (MAK064, SIGMA-ALDRICH^®^, St Louis, MO, USA), and catecholamine release (BA E-6500R, LDN^®^, Nordhorn, Germany) by standard enzyme-linked immunosorbent assay (ELISA) kits according to the manufacturer’s instructions.

### Statistical Analysis

Data are presented as the mean ± standard deviation (SD). All statistical analyses were completed using GraphPad Prism 8.0. Repeated measures ANOVA or linear mixed models (LMM) were used to analyze all metabolic and blood data, with Bonferroni post hoc to determine differences between trials. The model included main effects for time (exercise workload) and vaccine (Trial A vs Trial B) and interaction (Time x Trial) effects. Paired sample T-tests were used to detect differences in predicted 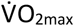 and time to ventilatory threshold between the trials performed during Visit 2 and Visit 3. Significance was set at *p* < .05

## Results

### COVID-19 vaccination is associated with increased physiological responses to graded cycling exercise in healthy individuals

To determine if COVID-19 vaccination is associated with changes in the physiological responses to exercise, we first of all compared pre and post vaccine exercise responses in the entire vaccinated cohort (n=12) regardless of SARS CoV-2 exposure status or vaccine type (Figure 1). Overall, the physiological responses to exercise were similar between trials but we did find significant interaction (Time x Trial) effects for oxygen uptake 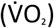, CO_2_ production 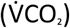, respiratory exchange ratio (RER), ventilation 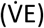, heart rate (HR), and serum adrenaline and noradrenaline levels, with each outcome measure indicating an increased metabolic demand of exercise after vaccination. Post-hoc analysis revealed that HR and noradrenaline levels were elevated at the 80% 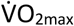 stage compared to the pre vaccine trial (p = 0.04 and 0.002 respectively). The RPE tended to be lower post vaccine at the 50% 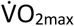 stage (p = 0.06) but not at the other exercise intensities. We found no pre-to-post vaccine differences for the ventilatory equivalents of oxygen uptake 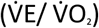, carbon dioxide production 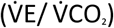, stroke volume (SV), cardiac output (CO), predicted 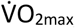, time to ventilatory threshold, serum lactate, and serum cortisol.

**Figure 1.**
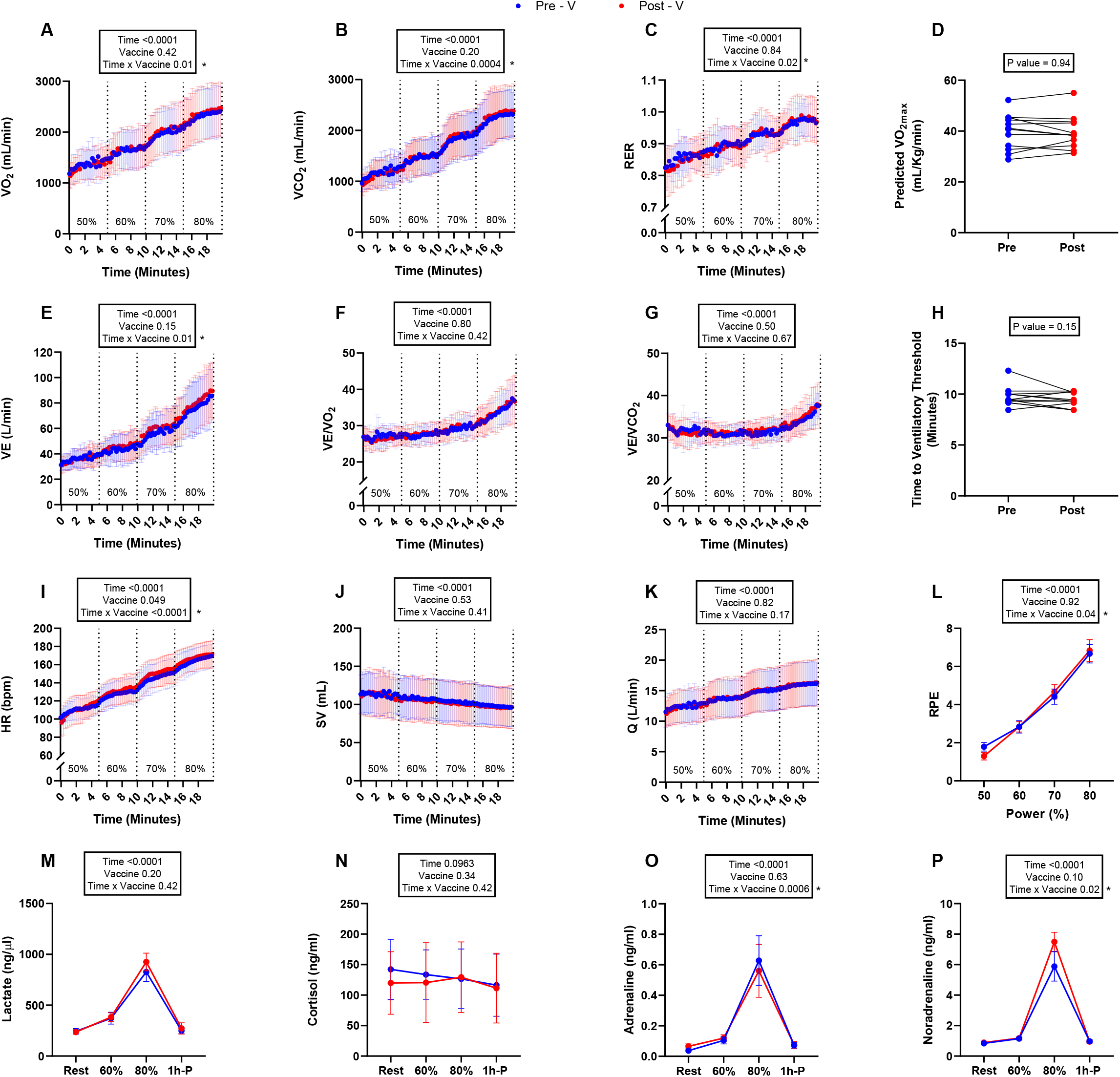
Metabolic and physiological responses to graded exercise pre (blue circle) and post (red circle) COVID-19 vaccination (n=12). (A) 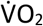, (B) 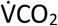, (C) RER, (D) Predicted 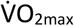, (E) VE, (F) 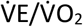, (G) 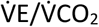, (H) Time to ventilatory threshold, (I) HR, (J) SV, (K) Q, (L)RPE, (M) Lactate, (N) Cortisol, (O) Adrenaline, and (P) Noradrenaline. Mean ± SD.

### The Pfizer mRNA COVID-19 vaccine increased heart rate and noradrenaline in response to graded cycling exercise in non-infected healthy individuals

To determine if these changes in the physiological responses to graded exercise after vaccination held true in our non-infected cohort who received the Pfizer mRNA vaccine (*n*=8), we found that the elevation in HR and noradrenaline response at the 80% 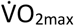 stage was still significant (*p* = 0.007 and 0.001, respectively), as well the interaction effect for RER elevated in response to exercise post-vaccine (Figure 2). We did not find differences in any other physiological endpoint post vaccine, although many of the interaction effects found to be significant in the entire vaccinated cohort were still trending in the non-infected Pfizer mRNA vaccine cohort (Figure 2).

**Figure 2.**
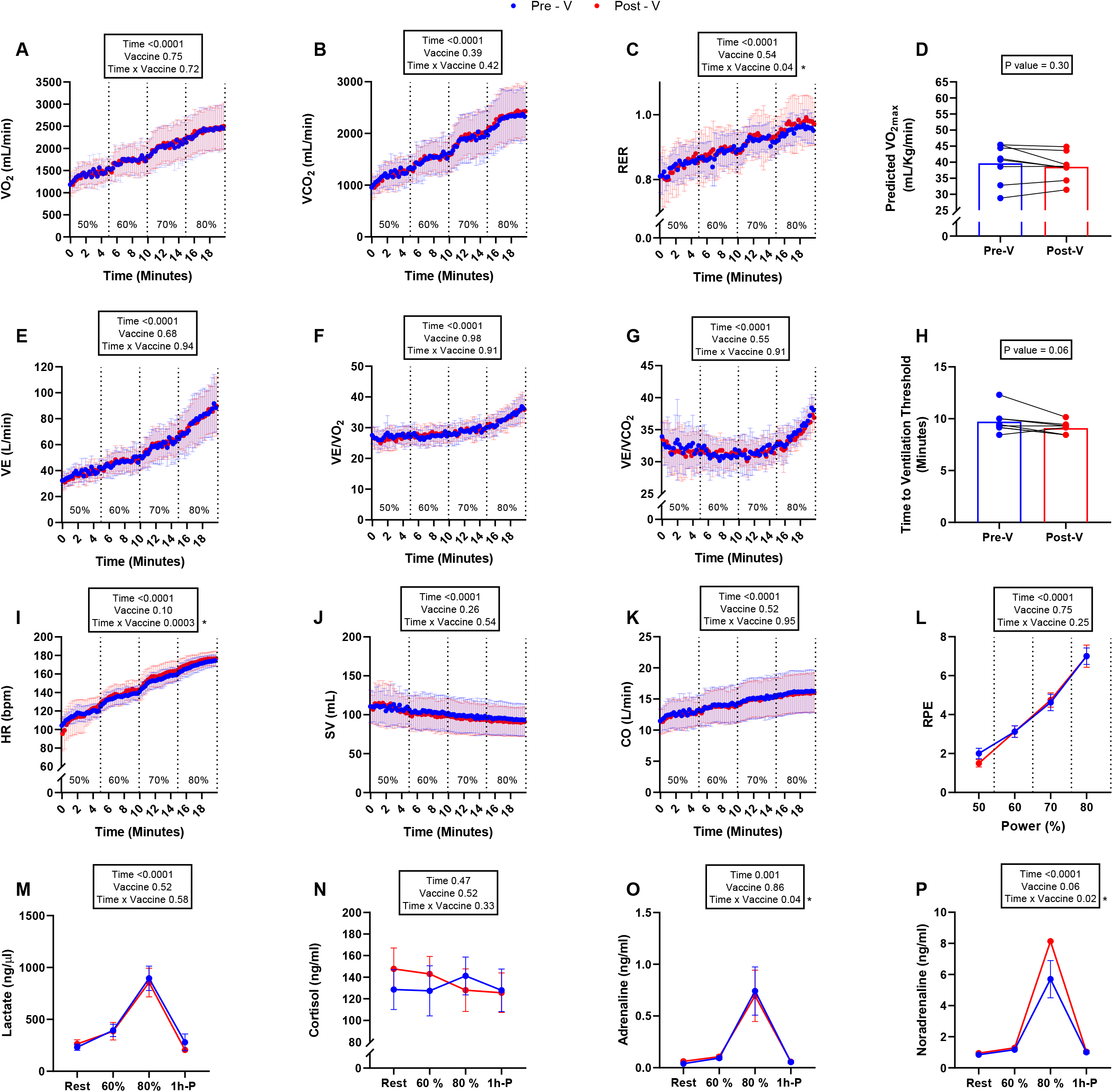
Metabolic and physiological responses to graded exercise pre (blue circle) and post (red circle) Pfizer mRNA Covid-19 vaccine in non-infected healthy individuals (n=8). (A) 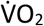, (B) 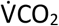, (C) RER, (D) Predicted 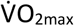, (E) VE, (F) 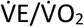, (G) 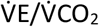, (H) Time to ventilatory threshold, (I) HR, (J) SV, (K) Q, (L)RPE, (M) Lactate, (N) Cortisol, (O) Adrenaline, and (P) Noradrenaline. Mean ± SD.

### COVID-19 vaccination is associated with improvements in the metabolic response to graded cycling exercise in previously infected healthy individuals

Due to emerging reports that COVID-19 vaccination is associated with improved symptomology following SARS-CoV-2 infection (12), we sought to determine if metabolic responses to graded exercise would be affected by the vaccine in those with natural immunity to the virus (Figure 3). In this small cohort of 4 participants who received either the Pfizer mRNA vaccine (n=1) or the Johnson&Johnson viral vector-based vaccine (n=3), we found that RER and adrenaline were lower after the vaccine at 80% of the 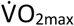 *(p* = 0.03, and 0.02 respectively) compared to the pre-vaccine trial. Serum lactate also followed a similar trend (*p* =0.07). No differences were found between the pre and post vaccine trials in this cohort for any other endpoint measures.

**Figure 3.**
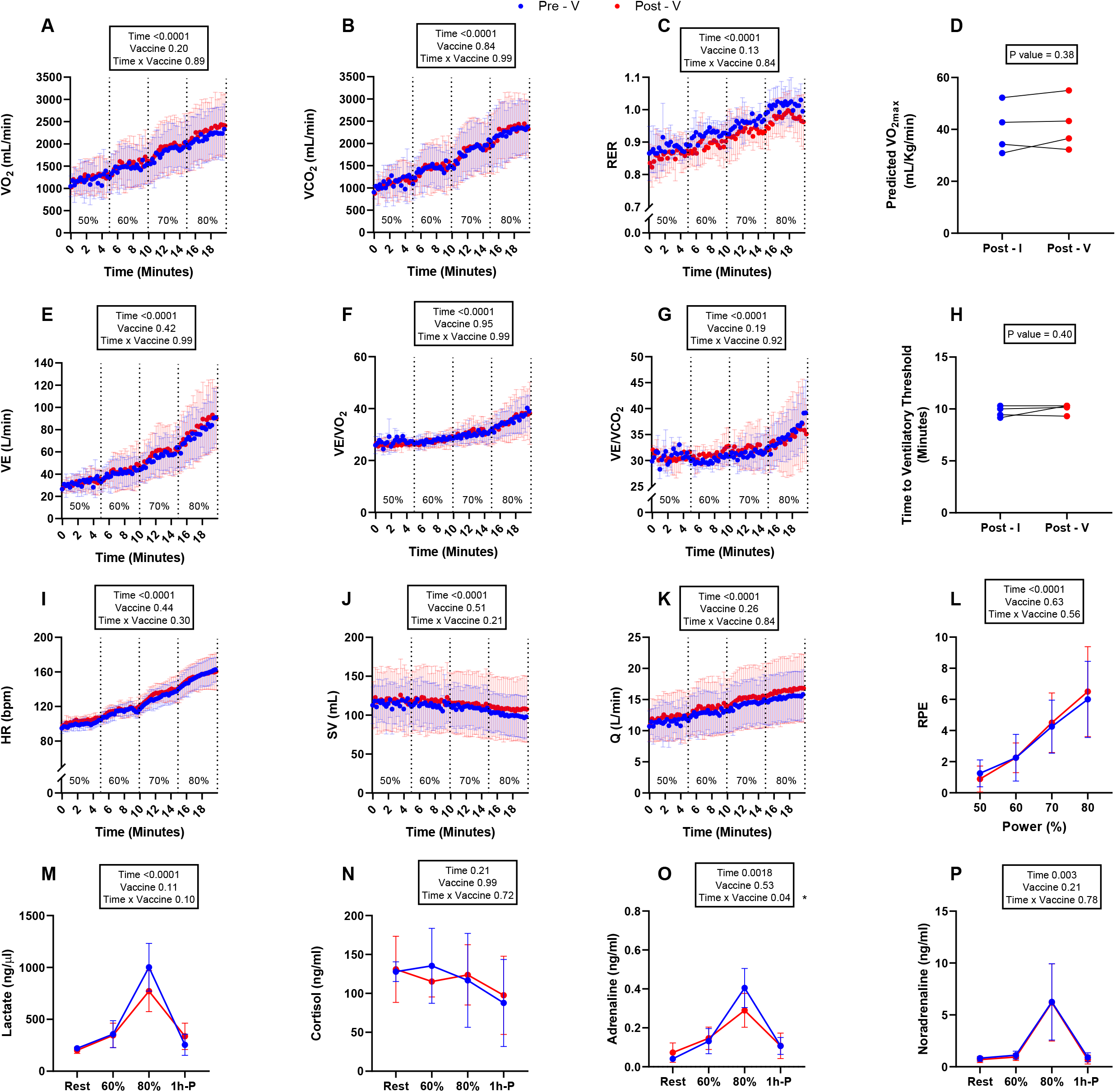
Metabolic and physiological responses to graded exercise pre (blue circle) and post (red circle) Covid-19 vaccine in previously infected healthy individuals (n=4). (A) 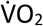, (B) 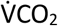, (C) RER, (D) Predicted 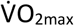, (E) VE, (F) 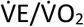, (G) 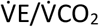, (H) Time to ventilatory threshold, (I) HR, (J) SV, (K) Q, (L)RPE, (M) Lactate, (N) Cortisol, (O) Adrenaline, and (P) Noradrenaline. Mean ± SD.

**Figure 4.**
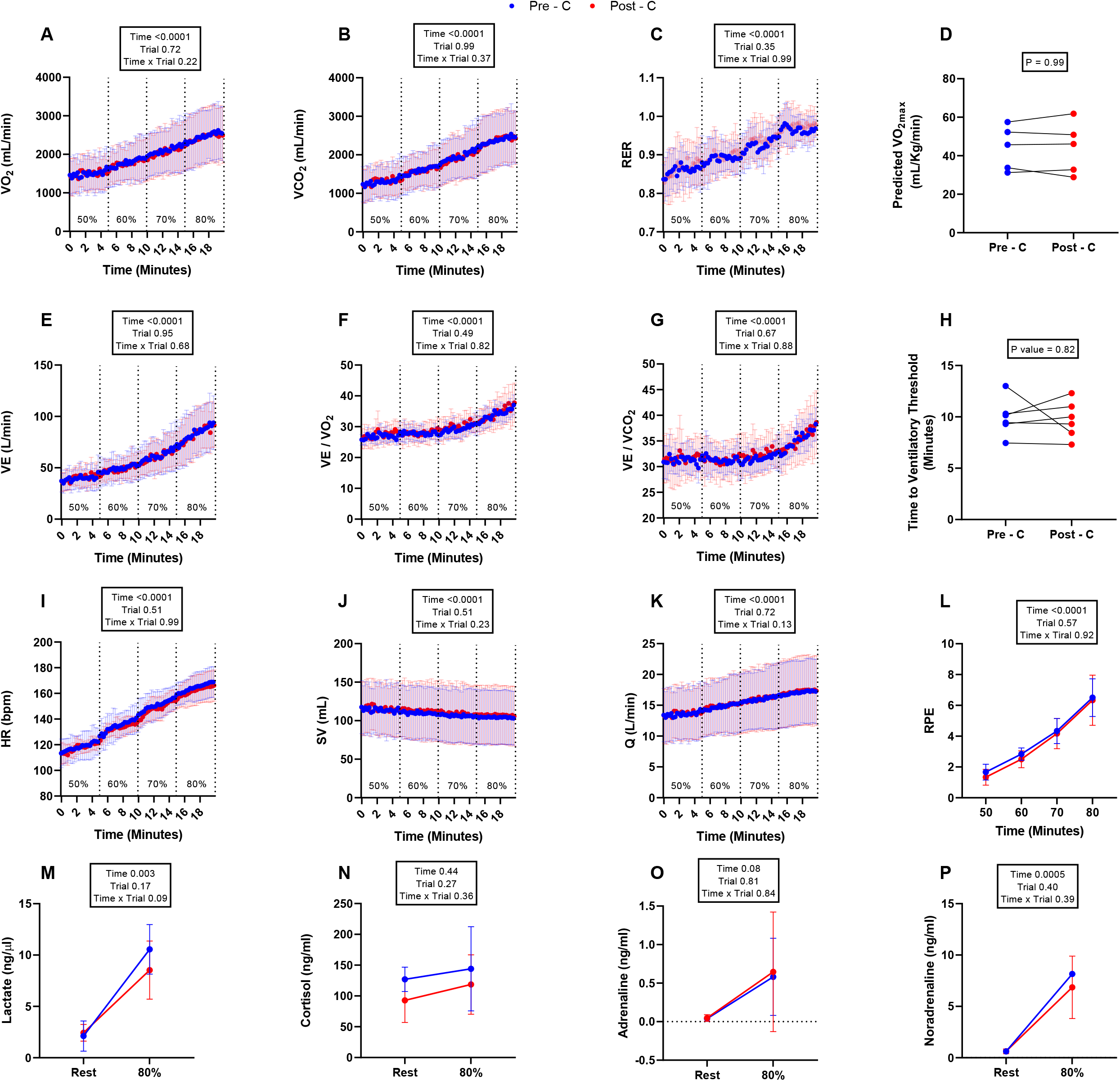
Metabolic and physiological responses to graded exercise in two separate trials without COVID-19 vaccination. Trial 1 (Blue circle) Trial 2 (red circle) (n=6). (A) 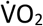, (B) 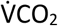, (C) RER, (D) Predicted 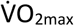, (E) VE, (F) 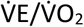, (G) 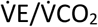, (H) Time to ventilatory threshold, (I) HR, (J) SV, (K) Q, (L)RPE, (M) Lactate, (N) Cortisol, (O) Adrenaline, and (P) Noradrenaline. Mean ± SD.

### Repeat exercise testing elicits identical responses to graded exercise in control participants that did not receive COVID-19 vaccination

As this study was not randomized, we decided to include data collected from a parallel study being performed in our laboratory whereby two bouts of graded exercise were performed by healthy participants ~5-weeks apart (i.e., similar to the time elapsed between Visit 2 and Visit 3 in the present study) without receiving a vaccine (Figure 3). All participants in this group were found to be seronegative for SARS-CoV-2 at the time of testing (Visit 2 and Visit 3). The exercise bouts performed by these control participants were identical to the vaccinated cohorts described here with the exception of the aforementioned less frequent blood sampling schedule. Overall, we found that the physiological responses to graded exercise were identical between these two trials and found no main or interaction effects that were identified in our vaccinated cohorts. There was, however, a trend for serum lactate to be lower on the second trial at 80% of the 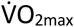 (p = 0.09), which was similar to what we found with our previously infected cohort in the post vaccine trial (Figure 2).

## Discussion

Vaccination is strongly recommended to safeguard athletes from infection during training and competition (7). Several major sporting events (e.g., UEFA European and Copa America Soccer Championships, Tokyo Olympic Games) have been held during the COVID-19 pandemic, increasing the risk of SARS-CoV-2 infection for non-vaccinated athletes. While both vaccination (13) and natural immunity (e.g. from prior infection) (14) can protect against COVID-19 disease, non-vaccinated athletes are at an increased risk of contracting SARS-CoV-2, which could result in them missing major sporting events and initiating isolation protocols for other athletes they were in close contact with. Despite this risk, anecdotal reports have emerged of athletes refusing the COVID-19 vaccine due to perceived negative impacts it may have on both their health and performance.

This is the first study, to our knowledge, to report on physiological responses to exercise before and after COVID-19 vaccination. We found that recent COVID-19 vaccination in a group of physically active healthy individuals was associated with an increased physiological demand to fixed intensity graded cycling exercise, evident by elevations in oxygen uptake, CO2 production, respiratory exchange ratio, ventilation, heart rate, rating of perceived exertion and plasma noradrenaline levels in the exercise trial performed 2-3 weeks after vaccination. We did not find changes in these parameters in a cohort of control participants who completed identical bouts of exercise several weeks apart without receiving a vaccine. Although it is possible that these effects are due to reduced physical activity levels after vaccination (e.g., due to symptoms of vaccinosis), we deem a detraining effect unlikely as, despite reporting many of the common symptoms associated with COVID-19 vaccination, our participants did not report significant changes to their physical activity levels during the study period. While this indicates that the physiological demands to defined intensity exercise is greater after COVID-19 vaccination, we should emphasize that these changes were small in magnitude and that other key measures of metabolic load and exercise capacity (e.g., blood lactate, plasma adrenaline, plasma cortisol, predicted 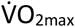 and ventilatory threshold) were unaffected by vaccination. As such, COVID-19 vaccination does not appear to evoke striking changes in aerobic exercise capacity or physiological responses to graded exercise in physically active healthy people. Nevertheless, it is possible that these small changes in the physiological response to graded exercise could impact athletic performance at the elite level and this warrants further investigation. The mechanisms by which COVID-19 vaccination might increase the physiological demands to graded exercise in healthy people are not known, although it would be expected that many of the metabolic shifts seen in the post-vaccine trial could be due to the heightened noradrenaline response to exercise that was concomitantly observed.

When analyzing a subset of our cohort who had never been exposed to SARS-CoV-2 and received two doses of the Pfizer mRNA vaccine, we found that many of these differences between the two exercise bouts remained (or were trending) significant. This indicates that these shifts in the physiological demands of exercise are most likely to occur in non-infected people at least 2-3 weeks after receiving the second dose of an mRNA vaccine. Conversely, we did not find increases in the physiological demands to exercise after vaccination in those who were previously infected with SARS-CoV-2. In fact, our data, albeit in a small number of participants, indicates that vaccination might even improve the metabolic response to exercise in those previously infected, evident by lower plasma adrenaline levels and a trend for reduced respiratory exchange ratio, and blood lactate levels. This could have ramifications for those experiencing symptoms of ‘long COVID’ as lowered exercise capacity is one of the hallmark features (15) and vaccination has been reported to ameliorate symptoms in these so-called ‘long-haulers’ (12). A potential confounder to these apparent divergent effects in the post-vaccine exercise response between previously infected and non-infected participants is the fact that most of our previously infected participants received a single dose human adenovirus-based vector vaccine (Johnson & Johnson) as opposed to the mRNA vaccine. We also cannot rule out a familiarization effect for the blood lactate response as we did observe a similar trend for lower serum lactate levels in the second trial performed by our control participants who did not receive a vaccine. Nevertheless, larger studies focused on how different vaccines affect physiological responses to exercise in individuals stratified by SARS-CoV-2 infection history and training status are warranted.

While providing the first report of physiological responses to graded exercise following COVID-19 vaccination, we acknowledge several study limitations. The small sample size may not only have prevented us from finding additional effects of vaccination on the exercise response, but also limited our ability to compare the response among infected and non-infected individuals who had received different types of COVID-19 vaccine. Further, we did not include an endpoint measure of exercise performance (e.g., cycling time trial or peak power test) and did not perform follow up assessments to determine the time course for these physiological responses to exercise to return to pre-vaccination levels. We purposefully tested our participants 2-3 weeks after vaccination as this is within the timeframe for neutralizing antibody production and SARS-CoV-2 T-cell detection (16), and because athletes are oftentimes vaccinated in close proximity to competition (7). As we did not administer the vaccines ourselves, we also had no control over the timing of the vaccine or the type of vaccine each individual received.

Despite these acknowledged limitations, we conclude that recent COVID-19 vaccination is associated with modest increases in the physiological demands to graded exercise, particularly in those receiving two doses of an mRNA vaccine and with no history of SARS-CoV-2 infection. These findings could have practical implications for athletes in regards to when COVID-19 vaccines are administered prior to and during major sporting events, particularly because booster shots or new vaccines may be required for continuous protection against SARS-CoV-2 and its evolving variants (17). We have also provided preliminary data that COVID-19 vaccination may actually be beneficial for the metabolic response to exercise in those previously infected with SARS-CoV-2, which could have implications, not just for athletes, but those suffering from long COVID syndrome. Future studies are required to determine if these small changes in the physiological demands to exercise after COVID-19 vaccination impacts athletic performance at the elite level, and whether or not the response is influenced by SARS-CoV-2 infection history and the type of vaccine administered.

## Summary box

- COVID-19 vaccination is associated with modest increases in the physiological demands to graded exercise, particularly in those with no history of SARS-CoV-2 infection.
- COVID-19 vaccination may improve the metabolic response to exercise in those with history of SARS-CoV-2 infection
- These findings could have implications for athletes in regard to when vaccines are administered prior to and during major sporting events

## Data Availability

Data can be made available on request by contacting the corresponding author.

